# Evaluating the Clinical Utility of a Brief Remote Speech Assessment for Screening and Triage of Speech Disorders

**DOI:** 10.64898/2026.11.16.26368191

**Authors:** Nicole V. Dickerson, Rene L. Utianski, Joseph R. Duffy, Heather M. Clark, Julie A. G. Stierwalt, Leland R. Bernard, Nathan P. Staff, Hugo Botha

## Abstract

**Purpose:** Speech abnormalities can often be early or clinically important manifestations of neurologic disease, yet access to clinicians with specialized expertise required for comprehensive motor speech evaluation may limit timely assessment. Brief remote speech assessment could provide a scalable approach for identifying patients who warrant more comprehensive evaluation, but its utility across heterogeneous speech presentations remains unclear. This study evaluated the validity and clinical utility of the Mayo Speech Test (MST), a brief (∼5-minute) audio-recorded remote speech assessment, relative to a comprehensive in-person motor speech examination (∼1-hour). Specifically, we examined whether MST could support clinically meaningful screening and triage decisions by distinguishing normal from abnormal speech and identifying the presence of a speech disorder.

**Method:** One hundred participants completed a comprehensive in-person speech evaluation and a remote MST. Participants were excluded when the interval between assessments exceeded 90 days (n = 13) or either rater judged audio quality insufficient for diagnostic interpretation (n = 6), with overlapping exclusions resulting in a final analytic sample of 82 participants. Remote recordings were independently evaluated by two speech-language pathologists (SLPs) who were blinded to participant identity. In-person evaluation classifications served as the clinical reference standard and were grouped as normal (n = 33), dysphonia only (n = 5), or speech disorder (n = 43). One participant could not be classified. Performance was evaluated for two clinically motivated binary decisions: distinguishing normal from abnormal (such as dysphonia or speech disorder; Tier 0) and identifying normal speech or dysphonia only versus speech disorder (Tier 1). Sensitivity, specificity, predictive values, agreement, and error patterns were examined.

**Results:** Both remote raters demonstrated similar overall performance despite differing error profiles. For Tier 0 screening, Rater 1 demonstrated 93.6% sensitivity and 71.9% specificity for identifying abnormal voice/speech, whereas Rater 2 demonstrated 78.7% sensitivity and 93.9% specificity. For Tier 1 identification of speech disorder, sensitivity and specificity were 83.3% and 86.5%, respectively, for Rater 1 and 71.4% and 97.4% for Rater 2. Interrater agreement was 79.7% for Tier 0 (κ = 0.60) and 86.1% for Tier 1 (κ = 0.72). Missed speech disorders represented an important source of error, with differences between raters indicating distinct thresholds for assigning abnormality and speech-disorder classifications.

**Conclusions:** A brief remote speech assessment showed promising performance for broad clinical classification, particularly when used to address screening and triage questions rather than reproduce the diagnostic granularity of a comprehensive examination. Similar overall performance across raters masked meaningful sensitivity-specificity tradeoffs, highlighting the importance of examining error profiles in addition to aggregate accuracy. MST may therefore have clinical utility for identifying patients who warrant comprehensive motor speech evaluation, but should not be considered a substitute for a full diagnostic assessment.

## Introduction

Access to specialized speech-language pathology (SLP) services can be limited by clinician availability, geographic distance, mobility limitations, and the time and resources required for comprehensive evaluation (Morton et al., 2022; Rameau et al., 2023). These challenges are particularly relevant for patients with neurologic disease, for whom access to clinicians with specific expertise in motor speech disorders may be limited. Workforce data further underscores this concern. In a 2025 survey of speech-language pathologists working in health care settings, more than half reported that job openings exceeded available job seekers in their geographic area and practice setting, and 44.8% reported funded but unfilled SLP positions at their facilities (American Speech-Language-Hearing Association, 2025). As demand for SLP services continues to increase, scalable approaches that can efficiently identify patients requiring comprehensive specialist evaluation may help extend the reach of limited clinical resources (Rezaii & Estiri, 2026).

Efficient access to motor speech assessment may be especially important because changes in speech can be an early or prominent manifestation of neurologic disease. Motor speech disorders, including dysarthria and apraxia of speech (AOS), occur across a range of acquired and neurodegenerative conditions and may provide clinically meaningful information about underlying neurologic dysfunction (Duffy, 2012). In Parkinson’s disease, for example, changes in voice and speech may occur early in the disease course and have been investigated as potential markers of disease presence and progression (Ma et al., 2020). In amyotrophic lateral sclerosis (ALS), physiologic changes across speech subsystems may be detectable before overt declines in speech intelligibility (Ball et al., 2002; Green et al., 2013, Rong et al., 2015). Speech impairment can also be the presenting or predominant manifestation in neurodegenerative disease, as demonstrated in primary progressive apraxia of speech (Duffy et al., 2021). Collectively, these findings highlight speech as a potentially sensitive behavioral window into neurologic function and underscore the value of timely identification of motor speech abnormalities.

Comprehensive motor speech evaluation and diagnosis, however, requires considerable clinical expertise and typically incorporates a series of speech tasks, perceptual judgments, examination of the speech mechanism, and consideration of the pattern and severity of abnormalities across speech subsystems (Duffy, 2012, Green et al., 2013). Such evaluations may be difficult to deploy broadly as an initial point of contact, particularly when the primary clinical question is whether a patient demonstrates findings that warrant specialist evaluation. A brief remote assessment capable of reliably distinguishing individuals with and without clinically meaningful speech abnormalities could therefore serve a different but complementary purpose – screening and triage rather than definitive diagnosis.

Telepractice and remote assessment have become increasingly feasible approaches for extending speech-language pathology services beyond traditional in-person settings (Theodoros, 2011; Weidner & Lowman, 2020). Early investigations demonstrated that assessment of dysarthria via telehealth was feasible and that several measures of severity, intelligibility, and perceptual speech characteristics could show acceptable agreement with face-to-face assessment, as well as high inter- and intra-rater reliability (Hill et al., 2006; Hill et al., 2009). Subsequent work has nevertheless highlighted important challenges, including the effects of recording equipment, compression, microphone placement, and internet connectivity on perceptual and acoustic speech measures (Dahl et al., 2021; Sevitz et al., 2021; Tran et al., 2026; Weerathunge et al., 2021).

The existing literature has frequently focused on specific clinical populations, individual acoustic or perceptual measures, or the remote administration of assessment protocols that largely mirror those used during a full comprehensive in-person evaluation (Ma et al., 2025; Migliorelli et al., 2023; Nunes et al., 2024; Theodoros et al., 2003). Considerably less is known about whether a brief, remotely obtained speech sample can provide sufficient information to support clinically useful motor speech screening and triage when compared with a comprehensive in-person evaluation, particularly across a heterogeneous population representing a range of neurologic diagnoses and speech presentations. This distinction is important because the value of a brief remote examination may not depend on its ability to reproduce the diagnostic granularity of a comprehensive motor speech evaluation. Instead, its greatest utility may lie in answering broader clinical questions such as: Does this patient’s speech appear abnormal? Is there evidence of a motor speech disorder? Does the patient warrant a more comprehensive evaluation? A remote tool that performs adequately at these levels could facilitate triage and prioritization while reserving more resource-intensive comprehensive examinations for patients most likely to benefit from them.

Mayo Speech Test (MST) is a brief remote speech assessment that can be captured in approximately five minutes and includes speech tasks intended to elicit clinically relevant features of speech production. In contrast, a comprehensive in-person motor speech evaluation may require an hour or longer and provides substantially greater diagnostic detail and information pertinent to clinical management recommendations. The purpose of the present study was, therefore, to evaluate the validity and clinical utility of MST relative to a comprehensive in-person motor speech examination. Rather than evaluating MST solely on its ability to reproduce a specific diagnostic classification, we examined its performance across clinically motivated levels of triage, including its ability to distinguish normal from abnormal voice/speech and to identify the presence of a speech disorder. We additionally examined interrater agreement and patterns of misclassification to characterize how clinician-related differences in decision thresholds may influence the performance of a brief remote assessment.

## Methods

### Participants

Participants were recruited as part of MST MIND-Mayo Speech Test: MIND (Monitoring Indicators of Neurologic Health and Disease) at Mayo Clinic in Rochester, Minnesota. The study was approved by the Mayo Clinic Institutional Review Board (IRB# 22-002430). The initial sample consisted of 100 consecutive participants who completed both a comprehensive in-person motor speech evaluation and a brief remote speech assessment using the Mayo Speech Test (MST). The sample was selected to include 50 consecutive participants whose in-person examination was completed by Rater 1 and 50 whose in-person examination was completed by Rater 2. Participants were excluded from the primary analyses if more than 90 days elapsed between the in-person and remote assessments or if either remote rater judged the remote recording as having insufficient audio quality to support clinical judgments. Thirteen participants exceeded the 90-day interval, and six had unusable audio recordings. Because one participant met both exclusion criteria, 18 unique participants were excluded, resulting in a final analytic sample of 82 participants. Of these, 41 had undergone the comprehensive in-person evaluation with Rater 1 and 41 with Rater 2. The median interval between the in-person evaluation and the remote recording was 21 days (IQR 12–34, range 0–82) and did not differ by in-person rater (median 21 days for each). Remote recordings were obtained after the in-person evaluation for 73 participants, on the same day for 1, and before it for 8.

The final cohort (Table 1) included 40 male and 42 female participants, ranging in age from 31 to 81 years (mean = 64 years, SD – 11.2). Most participants were White (n = 77, 93.9%) and not Hispanic or Latino (n = 76, 92.7%). Participants represented a heterogeneous clinical population (Figure 1): 46 (56%) had a degenerative/progressive neurologic diagnosis, 13 (16%) tremor or dystonia, 7 (9%) a functional neurologic disorder, and 6 (7%) other neurologic conditions (demyelinating, n = 2, n =1 for each of vascular, tumor, nonsurgical trauma, and compressive cranial neuropathy), while 10 (12%) had no neurologic diagnosis. The comprehensive in-person evaluations identified a range of speech abnormalities, including flaccid, spastic, ataxic, hypokinetic, hyperkinetic, unilateral UMN, and mixed dysarthria; AOS; functional speech disorder; other speech disorders (speech disruption due to structural change and neurogenic stuttering, n = 1 each; and isolated dysphonia (Figure 2). Because more than one diagnosis could be recorded per participant, and dysarthria types contributing to a mixed dysarthria were recorded individually, counts across categories exceed the number of participants with a speech disturbance.

**Figure 1.**
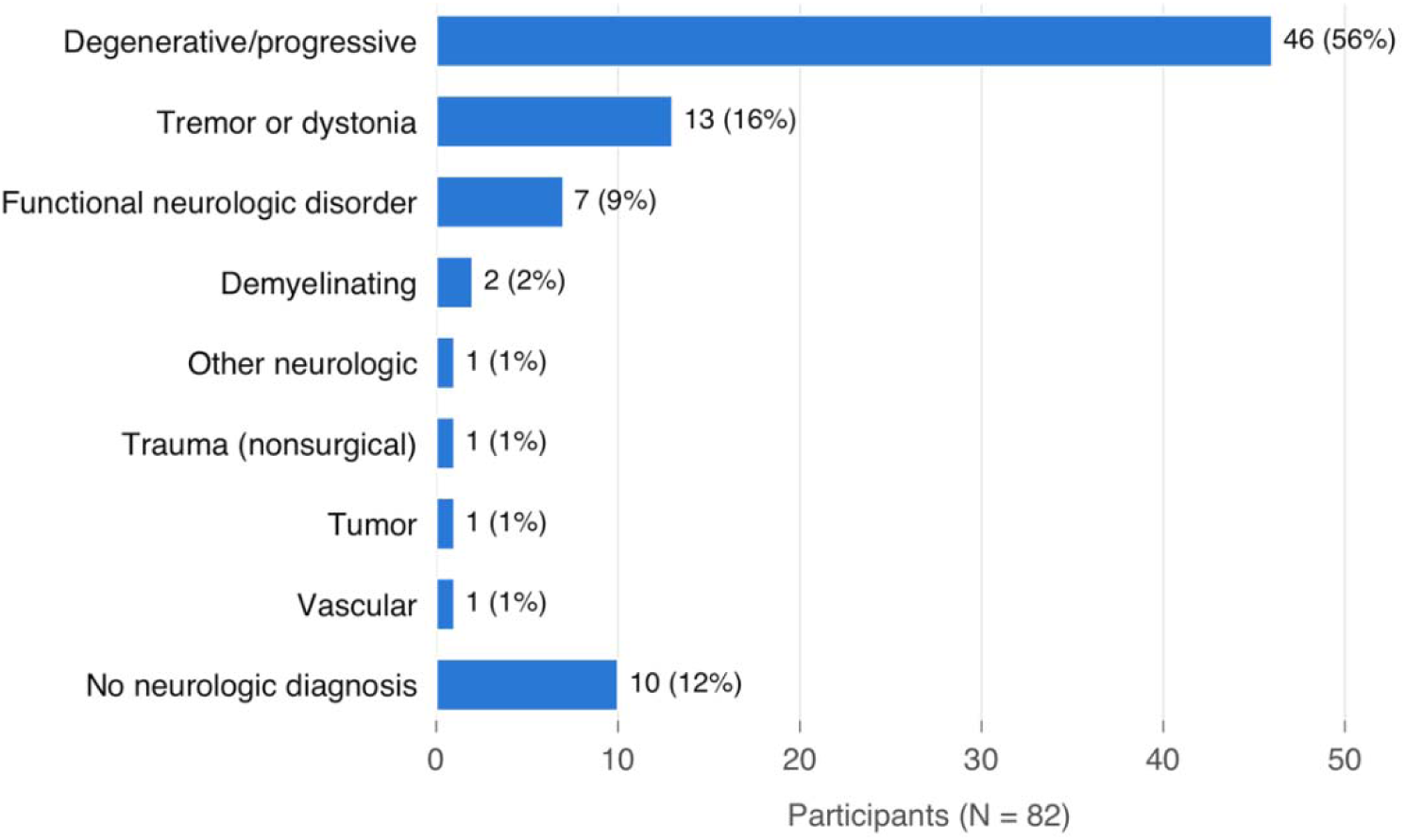
Neurologic diagnoses in the analytic cohort (N = 82)

**Figure 2.**
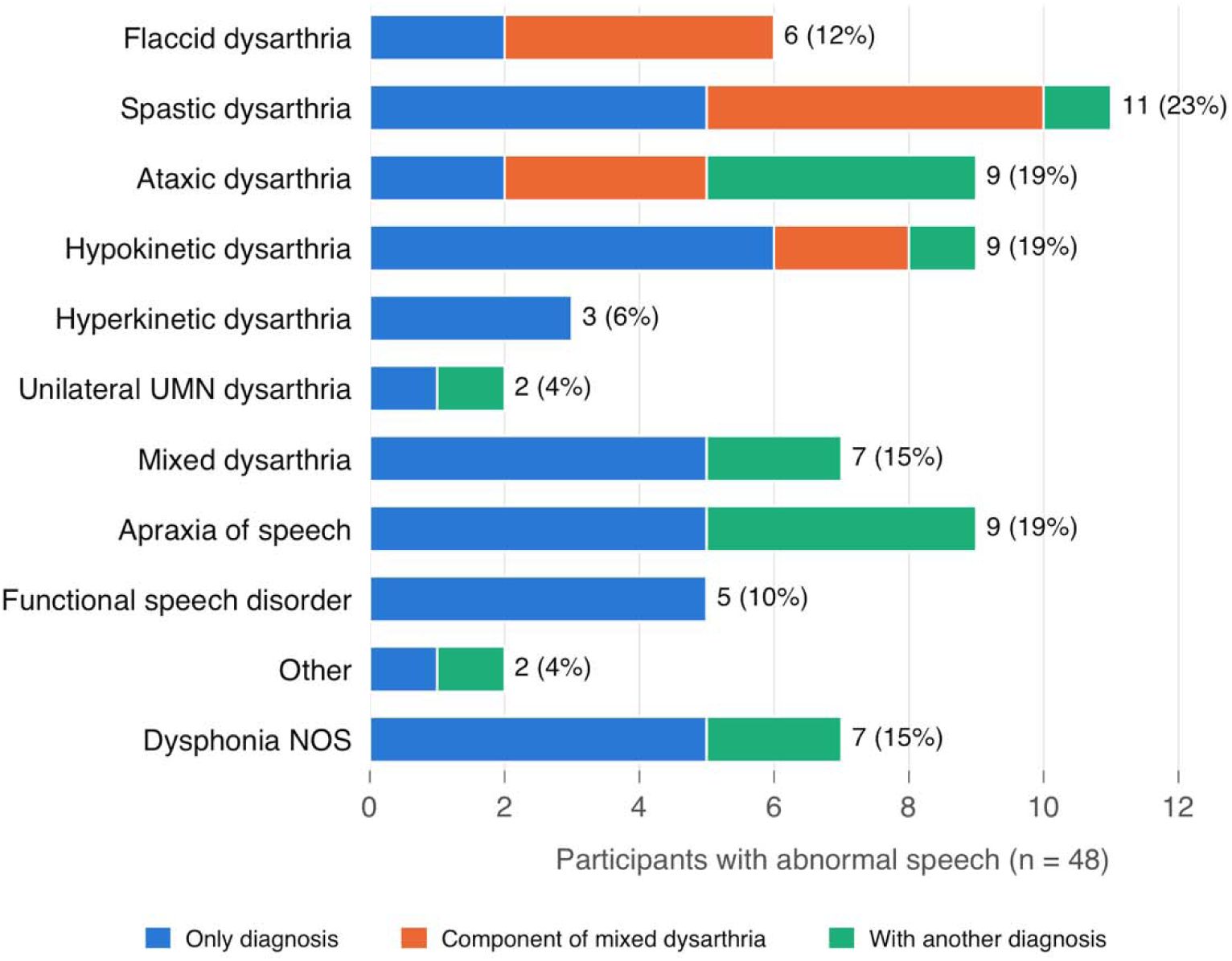
Reference-standard speech diagnoses among participants with abnormal speech (n = 48). Participants could receive more than one diagnosis. Each bar shows whether the diagnosis was the participant’s only diagnosis, a component type of a mixed dysarthria, or co-occurred with another speech diagnosis (e.g., AOS, dysphonia).

**Table 1.** Participant characteristics of the analytic cohort (N = 82)

| **Characteristic** | **Overall (N = 82)** | **In-person: Rater 1 (n = 41)** | **In-person: Rater 2 (n = 41)** |
|---|---|---|---|
| **Age, years, mean (SD)** | 64.0 (11.2) | 63.2 (11.4) | 64.8 (11.1) |
| **Age, years, median [IQR]** | 65.5 [58–73] | 65 [57–73] | 68 [59–73] |
| **Age, years, range** | 31–81 | 36–81 | 31–81 |
| **Female, n (%)** | 42 (51.2) | 23 (56.1) | 19 (46.3) |
| **Male, n (%)** | 40 (48.8) | 18 (43.9) | 22 (53.7) |
| **Race: White** | 77 (93.9) | 40 (97.6) | 37 (90.2) |
| **Race: Black or African American** | 2 (2.4) | 0 | 2 (4.9) |
| **Race: Not disclosed / unknown** | 3 (3.7) | 1 (2.4) | 2 (4.9) |
| **Ethnicity: Not Hispanic or Latino** | 76 (92.7) | 38 (92.7) | 38 (92.7) |
| **Ethnicity: Hispanic or Latino** | 1 (1.2) | 1 (2.4) | 0 |
| **Ethnicity: Not disclosed / unknown / unable to provide** | 5 (6.1) | 2 (4.9) | 3 (7.3) |
| **Days between in-person and remote, median [IQR]** | 21 [12–34] | 21 [13–33] | 21 [12–37] |
| **Reference standard: normal** | 33 (40.2) |  |  |
| **Reference standard: dysphonia only** | 5 (6.1) |  |  |
| **Reference standard: speech disorder** | 43 (52.4) |  |  |
| **Reference standard: unable to determine** | 1 (1.2) |  |  |

### Comprehensive In-Person Motor Speech Evaluation

Participants underwent a comprehensive in-person motor speech evaluation conducted by one of two certified speech-language pathologists (Rater 1 and Rater 2). The raters have expertise in the assessment of neurologic speech disorders and their clinical practice involves differential diagnosis of acquired and degenerative speech and language disorders in the context of medical workups for both known and unknown etiologies of neurologic symptoms. Comprehensive evaluations typically lasted approximately one hour and served as the clinical reference standard against which remote MST classifications were compared. The in-person examination routinely included reviewing medical history, oral mechanism examination, and perceptual evaluation (including sustained phonation, alternating and sequential motion rates, passage reading, repetition, and conversational speech). Based on the complete clinical examination, the evaluating SLP determined whether speech was normal or abnormal and, when appropriate, assigned a motor speech diagnosis and associated clinical characteristics, in alignment with the canonical Mayo Clinic classification system (DAB, 1969a,b; DAB,1975; Duffy, 2012).

### Mayo Speech Test

MST is a brief remote speech assessment designed to obtain clinically informative speech samples in approximately five minutes (Botha et al., 2026). Participants completed alternating and sequential motion rates, vowel prolongation, counting, word repetition, sentence repetition, passage reading, and picture description. Speech samples were recorded remotely by participants using their preferred electronic device, such a smart phone, computer, or tablet. Of the initial 100 recordings, 94 were considered to have sufficient audio quality to permit annotation of speech features and a diagnostic opinion. Six recordings were excluded because at least one rater judged the audio to be of insufficient quality.

Each MST recording was independently evaluated by two SLPs. Remote ratings were completed with raters blinded to participant identity. Raters completed no calibration exercise before rating and were not informed of the intended clinical use of their ratings (e.g., screening versus diagnostic confirmation), so that ratings would reflect each clinician’s usual diagnostic threshold. Based on the MST recording, raters provided their clinical impression, including whether speech was normal or whether isolated dysphonia and/or a speech disorder was present. When a speech disorder was identified, raters assigned a specific speech diagnosis (i.e., AOS, dysarthria subtype, functional speech disorder, or other). Raters could also indicate when a diagnosis could not be determined from the available recording. For each evaluation, raters reported their confidence in their diagnostic impression as not confident, somewhat confident, or very confident. Remote ratings were completed at a median of 418 days after the in-person evaluation, and for 75 of 82 participants (91%), more than six months had elapsed.

### Classification Framework

For analysis, the research team grouped the raters’ individual clinical diagnoses into three broader diagnostic categories: (1) normal, (2) dysphonia only, and (3) speech disorder. Participants were classified as normal when no voice or speech disorder was identified; as dysphonia only when non-specific voice differences were identified in the absence of a speech disorder; and as speech disorder when dysarthria, AOS, functional speech, or other speech disorder was identified, with or without dysphonia. When a rating form contained internally inconsistent selections (e.g., selecting both normal and dysarthria as the diagnosis), these were resolved with the rater before analysis. The same classification procedure was applied to diagnoses obtained from the comprehensive in-person evaluation documentation in the medical record, allowing the remote MST ratings to be compared with the in-person clinical reference standard using a common classification framework.

Within the final analytic cohort, application of this classification framework to the comprehensive in-person clinical reference standard resulted in 33 participants classified as normal, 5 as dysphonia only, and 43 with a speech disorder (dysarthria and/or AOS, n = 37; functional speech disorder, n = 5; speech disruption due to structural change (e.g., partial glossectomy), n = 1). For one participant, a definitive classification could not be determined. “Unable to determine” responses were treated as not classifiable rather than as a separate diagnostic category and were excluded from analyses requiring a definitive classification.

**Figure 3:**
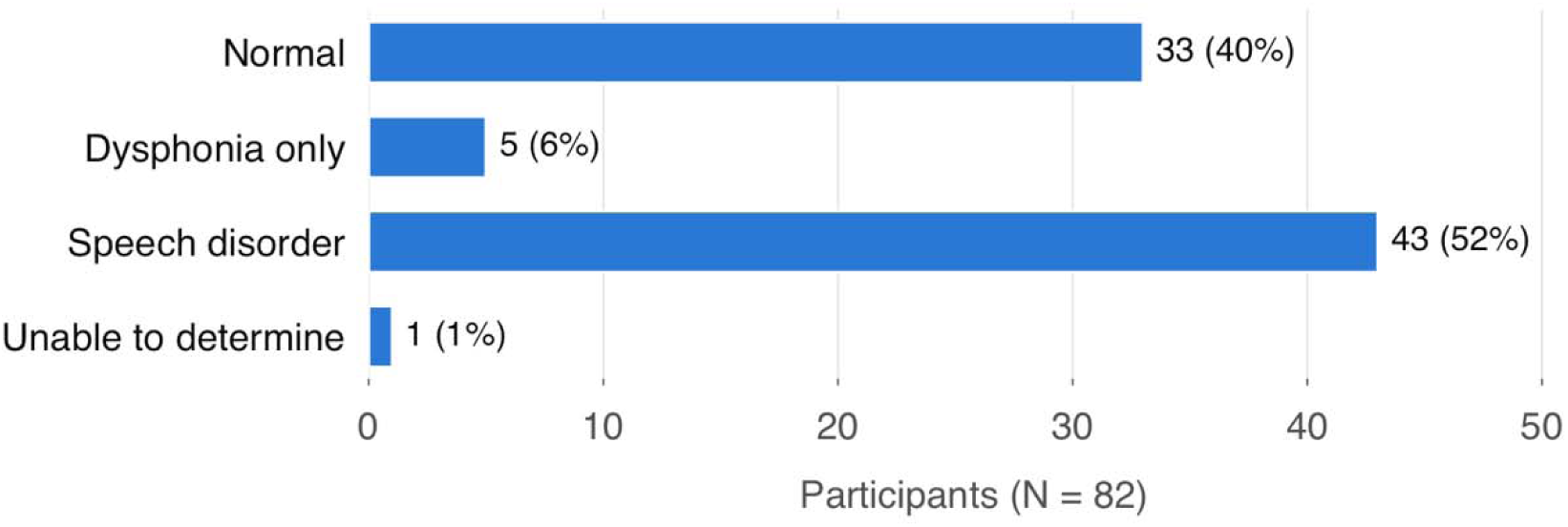
Reference-standard classification of the analytic cohort (N = 82).

Because MST is substantially shorter than a comprehensive motor speech evaluation, the primary analyses evaluated its utility for screening and triage rather than its ability to reproduce the full diagnostic granularity of the in-person examination. Using the diagnoses from the in-person reference standard, the three-category framework was therefore further collapsed into two clinically motivated binary classification tasks: Tier 0, normal versus abnormal speech, and Tier 1, normal speech or dysphonia only versus a clinically concerning speech disorder. Tier 0 evaluated whether MST allowed the raters to distinguish participants with normal speech from those with any identified abnormality; dysphonia-only and speech disorder classifications were combined into an “abnormal” category. This analysis represented an initial screening decision: *Does the speech sample contain evidence of an abnormality that may warrant further clinical evaluation?*

Tier 1 evaluated whether MST allowed the raters to identify the presence of a clinically concerning speech disorder. Participants classified as normal or dysphonia only were combined into a “no speech disorder” category and compared with participants with a speech disorder. This analysis represented the higher-stakes triage decision of primary interest: *Does the patient demonstrate evidence of a speech disorder warranting comprehensive motor speech evaluation to determine the specific nature of a speech disorder (e.g., dysarthria versus AOS vs functional speech disorder)?* This framework allowed evaluation of MST according to the clinical decisions a brief remote assessment might reasonably support rather than requiring exact reproduction of the diagnostic specificity available from a comprehensive examination.

### Statistical Analysis

Analyses evaluated both agreement between the two remote raters and performance of each remote rater relative to the comprehensive in-person clinical reference standard. For the three-category classification (normal, dysphonia only, and speech disorder), interrater agreement was quantified using percent agreement unweighted Cohen’s kappa, and linear-weighted kappa, treating the categories as ordered (normal < dysphonia only < speech disorder). Binary interrater agreement for the Tier 0 and Tier 1 classifications was quantified using percent agreement, Cohen’s kappa, and prevalence-adjusted bias-adjusted kappa (PABAK). Ninety-five percent confidence intervals for agreement statistics were estimated by percentile bootstrap (2,000 resamples of participants).

For comparisons with the clinical reference standard, diagnostic performance was characterized using sensitivity, specificity, positive predictive value (PPV), and negative predictive value (NPV). Exact (Clopper-Pearson) 95% confidence intervals were computed for sensitivity and specificity. A few cases in which either the reference-standard or corresponding remote classification was “unable to determine” were excluded from analyses requiring a definitive classification (reference standard, n = 1; Rater 1, n = 2; Rater 2, n = 1). Remote ‘unable to determine’ ratings were not scored as normal or absent, because in clinical deployment such cases would be referred for in-person evaluation rather than treated as negative. Consequently, 79 ratings were available for Rater 1 and 80 for Rater 2 for comparisons with the reference standard, and the participants with a reference-standard speech disorder included in each rater’s analysis (n = 42 each) differed by one participant. Interrater analyses were restricted to the 79 participants for whom both remote raters provided classifiable responses.

Error patterns were further characterized for the Tier 1 analysis. A false negative (FN) was defined as a participant classified as diagnosed with a speech disorder during the comprehensive in-person evaluation but classified as having no speech disorder based on MST recordings. A false positive (FP) was defined as a participant without a speech disorder according to the clinical reference standard labeled with a speech disorder based on MST recordings. FN and FP cases were examined by rater to characterize differences in clinical decision thresholds. Rater confidence was examined in relation to classification accuracy and error type. Distributions of confidence ratings were compared between raters with the Stuart–Maxwell test of marginal homogeneity for paired ratings, and with an exact McNemar test for use of the ‘very confident’ category. Confidence was compared between correct and incorrect Tier 1 classifications with Fisher’s exact test. Because the two raters differed in their use of the confidence scale, we did not interpret this as a uniform measure of diagnostic certainty across clinicians. All statistical analyses were conducted using R version 4.4.0 (R Core Team, 2024) using base functions.

## Results

### Interrater Agreement

Interrater analyses included 79 participants for whom both remote raters provided classifiable ratings. For the three-group classification of normal, dysphonia only, and MSD, the two remote raters agreed in 75.9% of cases (unweighted κ = 0.61, 95% CI 0.47-0.75; linear-weighted κ = 0.66, 95% CI 0.52-0.79). Agreement increased when diagnoses were collapsed into the clinically motivated binary classifications. For Tier 0 (normal vs. abnormal), percent agreement was 79.7% (κ = 0.60; PABAK = 0.59). For Tier 1 (speech disorder vs. no speech disorder), agreement was 86.1% (κ = 0.72; PABAK = 0.72).

**Table 2.**
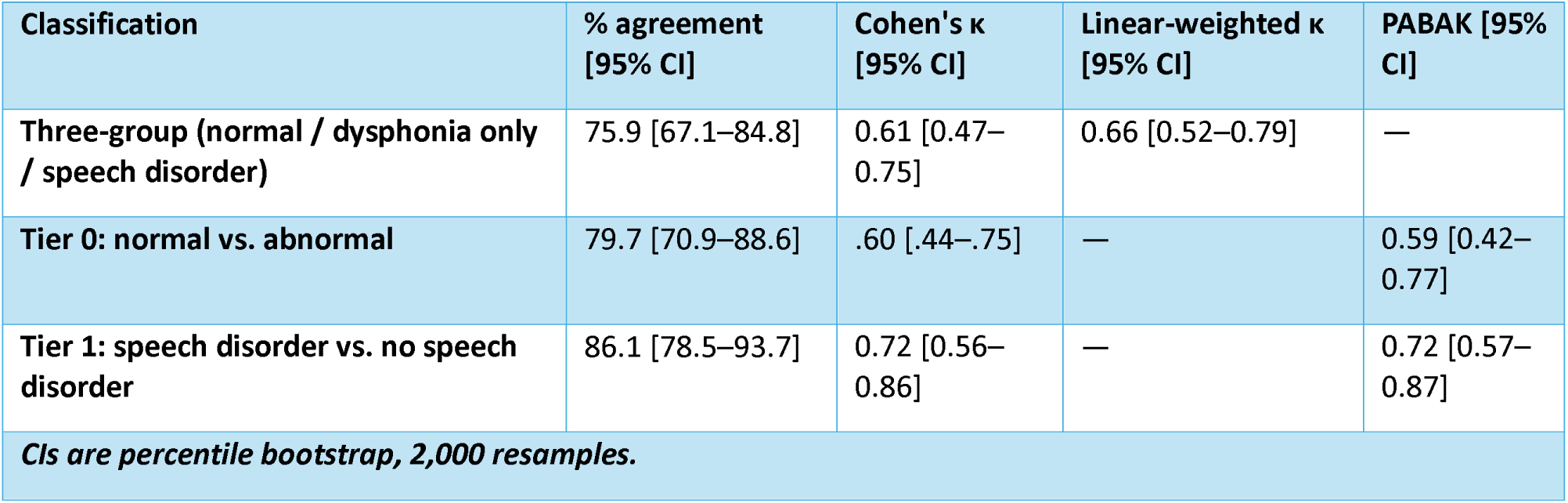
Interrater agreement between remote raters (n = 79)

### Tier 0: Normal Versus Abnormal Speech

Tier 0 evaluated the ability to identify abnormal speech, with dysphonia-only and MSD classifications combined into the abnormal category. Rater 1 classified 67 of 79 cases (84.8%) consistent with the in-person reference ratings. Sensitivity for detecting abnormal speech was 93.6% (95% CI 82.5–98.7), and specificity was 71.9% (53.3–86.3); PPV and NPV were 83.0% and 88.5%, respectively. Rater 1 had 3 false negatives and 9 false positives, relative to the reference standard.

Rater 2 classified 68 of 80 cases (85.0%) consistent with the in-person reference ratings. Sensitivity for detecting abnormal speech was 78.7% (95% CI 64.3–89.3), and specificity was 93.9% (79.8–99.3); PPV and NPV were 94.9% and 75.6%, respectively. Rater 2 had 10 false negatives and 2 false positives. Thus, although overall Tier 0 accuracy was similar between raters, the distribution of false-negative and false-positive classifications differed.

**Table 3.** Remote classification accuracy versus the in-person reference standard.

| Rater | Tier | n | TP | FP | TN | FN | Sensitivity % [95% CI] | Specificity % [95% CI] | PPV % | NPV % | Accuracy % |
|---|---|---|---|---|---|---|---|---|---|---|---|
| **Rater 1** | Tier 0 | 79 | 44 | 9 | 23 | 3 | 93.6 [82.5–98.7] | 71.9 [53.3–86.3] | 83.0 | 88.5 | 84.8 |
| **Rater 1** | Tier 1 | 79 | 35 | 5 | 32 | 7 | 83.3 [68.6–93.0] | 86.5 [71.2–95.5] | 87.5 | 82.1 | 84.8 |
| **Rater 2** | Tier 0 | 80 | 37 | 2 | 31 | 10 | 78.7 [64.3–89.3] | 93.9 [79.8–99.3] | 94.9 | 75.6 | 85.0 |
| **Rater 2** | Tier 1 | 80 | 30 | 1 | 37 | 12 | 71.4 [55.4–84.3] | 97.4 [86.2–99.9] | 96.8 | 75.5 | 83.8 |

*CIs are exact (Clopper–Pearson). Tier 0 positive = abnormal (dysphonia only or speech disorder). Tier 1 positive = speech disorder.*

### Tier 1: Speech Disorder Versus No Speech Disorder

Tier 1 evaluated the ability to identify the presence of a speech disorder, with normal and dysphonia-only classifications combined into the no-speech disorder category. Rater 1 classified 67 of 79 cases (84.8%) concordant with the in-person reference standard. Sensitivity for detecting a speech disorder was 83.3% (95% CI 68.6–93.0), and specificity was 86.5% (71.2–95.5); PPV and NPV were 87.5% and 82.1%, respectively. Of the 42 participants with a reference-standard speech disorder, Rater 1 identified 35 and had seven false negatives; there were five false positives.

Rater 2 classified 66 of 80 cases (82.5%) concordant with the in-person reference standard. Sensitivity for detecting a speech disorder was 71.4% (95% CI 55.4–84.3), whereas specificity was 97.4% (86.2–99.9); PPV and NPV were 96.8% and 75.5%, respectively. Of the 42 participants with a reference-standard speech disorder, Rater 2 identified 30 and had 12 false negatives, with one false positive. As observed for Tier 0, overall Tier 1 accuracy was similar between raters despite differences in the distribution of errors.

### Error Patterns and Rater Confidence

Across both raters, Tier 1 errors consisted of 19 false negatives (13 participants, 6 missed by both raters) and 6 false positives (5 participants, 1 incorrectly labeled by both raters). Among the 19 false negatives, 11 (57.9%) were rated with *somewhat confident* diagnostic confidence and 7 (36.8%) were rated *very confident*; only 1 (5.3%) was rated *not confident*. In contrast, none of the six false positives were accompanied by a *very confident* rating; four (66.7%) were rated *somewhat confident* and two (33.3%) were rated *not confident*. Thus, although false positives were not associated with high diagnostic confidence, several false negatives occurred despite the rater reporting high confidence in the remote diagnostic impression.

Missed speech disorders (false negatives) were concentrated in hypokinetic dysarthria. Of the 9 participants with reference-standard hypokinetic dysarthria, each rater identified 4 (44.4%), compared with 31 of 33 (93.9%; Fisher’s exact p = 0.003) and 26 of 33 (78.8%; p = 0.09) participants with other speech disorders for Raters 1 and 2, respectively. The same 5 participants with hypokinetic dysarthria were missed by both raters. Rater 1 classified 3 of these as dysphonia only and 2 as normal, whereas Rater 2 classified all 5 as normal, each with a very confident rating. Other missed diagnoses were AOS (Rater 1, n = 2; Rater 2, n = 3), ataxic dysarthria (n = 1; n = 2), and, for Rater 2 only, flaccid, hyperkinetic, and unilateral UMN dysarthria, functional speech disorder, and structural speech disruption (n = 1 each). Of note, these diagnostic counts overlap, e.g., one participant had hypokinetic dysarthria plus AOS. Across classifiable remote ratings, Rater 1 characterized 22.8% of ratings as not confident, 49.4% as somewhat confident, and 27.8% as very confident, whereas Rater 2 characterized 3.8% as not confident, 37.5% as somewhat confident, and 58.8% as very confident. Differences were also apparent among participants with a reference-standard speech disorder: Rater 1 reported being very confident for 6 of 42 (14.3%) cases, compared with 19 of 42 (45.2%) for Rater 2. Among the 78 participants classifiable by the reference standard and both raters, the distribution of confidence ratings differed between raters (Stuart–Maxwell χ²(2) = 23.3, p < 0.001). Rater 2 alone gave a very confident rating for 29 participants, compared with 5 for Rater 1 alone (exact McNemar p < 0.001). Within each rater, very confident ratings were less frequent for incorrect than correct Tier 1 classifications (Rater 1: 1 of 12 vs. 21 of 67; Rater 2: 6 of 13 vs. 41 of 67), but these differences were not statistically significant (Fisher’s exact p = 0.16 and 0.37)

**Table 4.**
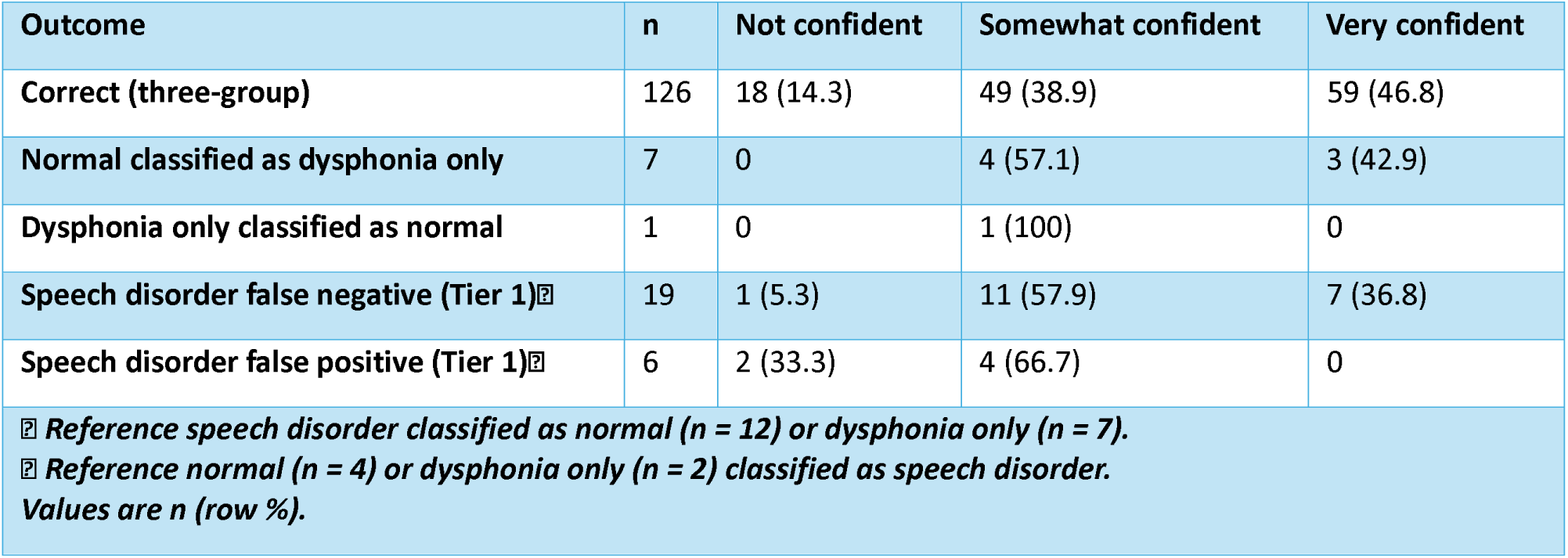
Diagnostic confidence by classification outcome (pooled ratings, n = 159)

| Outcome | n | Not confident | Somewhat confident | Very confident |
|---|---|---|---|---|
| **Correct (three-group)** | 126 | 18 (14.3) | 49 (38.9) | 59 (46.8) |
| **Normal classified as dysphonia only** | 7 | 0 | 4 (57.1) | 3 (42.9) |
| **Dysphonia only classified as normal** | 1 | 0 | 1 (100) | 0 |
| **Speech disorder false negative (Tier 1)[illegible]** | 19 | 1 (5.3) | 11 (57.9) | 7 (36.8) |
| **Speech disorder false positive (Tier 1)[illegible]** | 6 | 2 (33.3) | 4 (66.7) | 0 |

[illegible] *Reference speech disorder classified as normal (n = 12) or dysphonia only (n = 7).*
[illegible] *Reference normal (n = 4) or dysphonia only (n = 2) classified as speech disorder.*
*Values are n (row %).*

### Within- and Between-Rater Comparisons

To examine whether prior in-person clinical exposure to a participant influenced remote classification performance, additional analyses compared cases in which the remote MST was rated by the same clinician who conducted the comprehensive in-person examination (within-rater) with those rated by the other clinician (between-rater). For Tier 0, sensitivity for detecting abnormal speech was identical for the within- and between-rater comparisons (85.1% and 87.2), whereas specificity was 78.8% within-rater and 87.5% between-rater. For Tier 1, sensitivity for detecting a speech disorder was 81.0% within-rater and 73.8% between-rater, while specificity was 89.5% and 94.6%, respectively. There were 8 false negatives in the within-rater comparison and 11 in the between-rater comparison. Overall, performance was broadly comparable when remote ratings were compared with evaluations performed by the same versus the other clinician, although sensitivity for detecting speech disorders was slightly lower in the between-rater analysis.

**Table 5.** Accuracy when the remote rater did (within) or did not (between) perform the in-person evaluation.

| Comparison | Tier | n | Sensitivity % [95% CI] | Specificity % [95% CI] | FN |
|---|---|---|---|---|---|
| **Within-rater** | Tier 0 | 80 | 85.1 [71.7–93.8] | 78.8 [61.1–91.0] | 7 |
| **Within-rater** | Tier 1 | 80 | 81.0 [65.9–91.4] | 89.5 [75.2–97.1] | 8 |
| **Between-rater** | Tier 0 | 79 | 87.2 [74.3–95.2] | 87.5 [71.0–96.5] | 6 |
| **Between-rater** | Tier 1 | 79 | 73.8 [58.0–86.1] | 94.6 [81.8–99.3] | 11 |

## Discussion

The purpose of this study was to evaluate the validity and clinical utility of Mayo Speech Test (MST), a brief remote speech assessment, relative to a comprehensive in-person motor speech evaluation. Specifically, we examined whether approximately five minutes of remotely obtained audio-recorded speech could provide sufficient information to support clinically meaningful screening and triage decisions across a heterogeneous clinical population. Overall, the findings support the potential utility of MST for this purpose. Using MST recordings, two independent raters demonstrated approximately 85% accuracy for distinguishing normal from abnormal voice/speech and approximately 84% accuracy for identifying the presence or absence of a speech disorder. Interrater agreement was also higher for the clinically focused binary classification of speech disorder versus no speech disorder than for the more granular three-group classification of normal, dysphonia only, and speech disorder. Collectively, these findings suggest the clinical value of a brief remote audio-recorded assessment in identifying patients who may warrant further specialist evaluation.

An additional finding relevant to clinical implementation was the high proportion of remotely obtained recordings that were clinically interpretable. Of the 100 recordings initially obtained, only six were excluded because at least one rater judged the audio quality insufficient to support a diagnostic opinion, yielding a 94% rate of usable recordings. Three out of the six excluded cases were patients with hypokinetic dysarthria. Because recordings were obtained on participants’ own devices without a standardized mouth-to-mic distance, reduced loudness – a distinguishing feature of hypokinetic dysarthria - may be less apparent remotely, which may be relevant to the lower detection of hypokinetic dysarthria observed here. Nevertheless, the high proportion of usable recordings provides preliminary support for the feasibility of brief remote speech assessment, as clinical utility depends on reliably obtaining adequate speech samples outside of traditional in-person settings (Sevitz et al., 2021; Tran et al., 2026). The small number of unusable recordings also underscores the need for timely procedures to identify inadequate samples and obtain repeat recordings when necessary.

An important finding was that similar overall accuracy did not necessarily reflect similar classification behavior between clinicians. For Tier 0, Rater 1 demonstrated higher sensitivity for detecting abnormal voice/speech (93.6%) but lower specificity (71.9%), whereas Rater 2 demonstrated lower sensitivity (78.7%) but higher specificity (93.9%). A similar pattern emerged for Tier 1. Rater 1 identified 81.0% of participants with a speech disorder, with a specificity of 86.5%, whereas Rater 2 identified 71.4% of participants with a speech disorder but demonstrated a specificity of 97.4%. Consequently, Rater 1 produced fewer false negatives but more false positives, whereas Rater 2 produced fewer false positives but more false negatives.

These findings demonstrate that aggregate accuracy alone may provide an incomplete picture of the performance of a brief remote screening assessment. Importantly, raters were not instructed to approach the MST recordings with a specific clinical purpose (e.g., screening/ triage versus diagnostic assessment) and therefore were not directed toward a particular decision threshold. In a triage context, the balance between sensitivity and specificity is particularly important because failing to identify an individual with an MSD may delay referral for comprehensive evaluation, whereas an overcall is more likely to result in additional, perhaps unnecessary comprehensive assessment. Thus, calibration between raters, against an explicitly defined clinical purpose, could help establish shared decision thresholds and improve consistency in future applications of MST.

The pattern of Tier 1 errors further highlights this distinction. Across both raters, 19 remote ratings (of 13 participants; 6 missed by both raters) did not identify a speech disorder that was identified during the comprehensive in-person evaluation; in only six ratings (of 5 participants) was a speech disorder identified remotely that was not present during the clinical reference standard. Thus, missed identification of speech disorders represented the predominant clinically consequential outcome in this study. These false negative cases were concentrated in hypokinetic dysarthria: both raters missed the same five of nine participants with hypokinetic dysarthria, whereas detection of other speech disorders was higher (Rater 1, 93.9%; Rater 2, 78.8%). The brief nature of MST may contribute to these errors because subtle speech abnormalities may not be consistently apparent within a limited speech sample. A comprehensive examination provides substantially more opportunity to elicit and characterize abnormalities across tasks and speech subsystems. Accordingly, a negative finding on a brief remote assessment should not be interpreted as definitive evidence that a speech disorder is absent, particularly when other clinical information raises concerns. An expanded MST, with increased opportunities for connected speech, could be considered in the future with re-evaluation of concordance with conclusions from in-person testing.

The confidence findings provide additional insight into the nature of these errors. Most missed speech disorders occurred when raters reported being at least somewhat confident in their diagnostic impression, and seven of the 19 missed speech disorder ratings were accompanied by a rating of very confident. In contrast, none of the six false positives were rated as very confident. Confidence also differed considerably between raters: Rater 2 used the very confident category substantially more often than Rater 1. These findings suggest that confidence may capture aspects of an individual clinician’s certainty but should not be assumed to correspond directly to classification accuracy. Importantly, high confidence did not eliminate the possibility of a false-negative. If confidence is incorporated into future triage algorithms or clinical workflows, it may be most useful when interpreted alongside the diagnostic decision and other clinical information rather than as an independent indicator of the likelihood that classification is correct.

Interrater findings similarly support the use of broader classifications for remote screening. Agreement between the two remote raters was 75.9% for the three-group classification but increased to 86.1% when the clinical question was modified to the presence versus absence of a speech disorder. This pattern supports a potential role for MST during triage. Distinguishing among specific diagnostic categories requires a level of perceptual and diagnostic precision that may be difficult to achieve relaibility from a brief remote sample. In contrast, determining whether there is sufficient evidence of a speech disorder to warrant further evaluation represents a broader clinical decision and may be better suited to the amount of information available from MST.

The within- and between-rater analyses also provide useful context for interpreting agreement with a comprehensive evaluation. Tier 0 sensitivity was similar across within- and between-rater comparisons, while Tier 1 sensitivity was slightly higher within-rater than between-rater. Specificity was somewhat higher in the between-rater comparisons for both tiers. Although these analyses cannot completely exclude the possibility of participant recognition, the persistence of similar performance when the remote rater had not conducted the participant’s comprehensive evaluation suggests that prior clinical exposure alone is unlikely to account for the observed (dis)agreement.

Finally, our results should be interpreted in the context of what was a deliberately stringent test. The raters completed no calibration exercise and were not told whether their ratings were intended for screening, triage, or diagnostic confirmation. This was intentional, to avoid biasing them toward sensitivity or specificity, but it means each rater applied their own implicit decision threshold, consistent with the distinct sensitivity-specificity profiles observed. The results should therefore not be taken as a targeted assessment of the performance of MST in any single role, such as screening. Raters also had no control over the speech sample: participants recorded a fixed task set on their own devices, the tasks may not match each clinician’s own practice, and raters could not request repetitions or follow up with targeted tasks. Raters received no clinical context, such as a presenting complaint or referral question, whereas the in-person reference evaluation drew on the full medical record, patient reported history, and clinical examination. MST is also limited to audio, whereas the in-person evaluation incorporates visual and tactile information during the motor speech exam. Together with the interval between assessments and the imperfect reference standard discussed below, these conditions suggest that MST may perform better in a deployed workflow with a defined purpose, calibrated raters, and basic clinical context.

### Limitations and Future Directions

Several limitations should be considered when interpreting these findings. First, although remote MST recordings were restricted to those obtained within 90 days of the comprehensive in-person evaluation, clinical stability during this interval could not be confirmed. Participants may have experienced changes in neurologic status, speech function, or other factors affecting speech between assessments. Additionally, potentially relevant within-day factors were not controlled. For example, medication timing relative to assessment was not available for participants with Parkinson’s disease or parkinsonism (n = 6), and fluctuations associated with medication state, fatigue, or the variable presentation of some conditions (e.g., functional speech disorder) could have contributed to *real* differences between the in-person and remote speech presentations. Thus, some disagreement between assessments may reflect genuine changes or fluctuations in speech rather than differences in the ability of MST to capture motor speech abnormalities.

Second, the same two SLPs who rated the remote MST recordings also participated in the comprehensive in-person evaluations from which the clinical reference standard was derived. While this was by design, and remote ratings were completed without participant identifiers, it is possible that a rater could have recognized a participant from their voice or speech characteristics. This risk was likely reduced by the brief nature of the remote recordings and the time elapsed between the in-person evaluation and the remote rating (more than six months for 91% of participants) Additionally, performance remained generally similar when within-rater and between-rater comparisons were examined, suggesting that prior clinical exposure was unlikely to fully account for the observed agreement between MST and comprehensive evaluation. Nevertheless, future validation by independent raters with no prior clinical contact with participants would provide a more stringent assessment of performance.

Third, the comprehensive in-person evaluation served as the clinical reference standard rather than some sort of objective biological gold standard. Motor speech diagnosis relies in part on expert perceptual judgment, and some degree of clinician variability is therefore expected (Pernon et al., 2022). This consideration is particularly relevant given the rater-specific patterns observed in the present study. Although overall classification accuracy was similar between raters, their sensitivity and specificity differed, particularly for identification of motor speech disorders. These findings suggest that individual clinicians may apply different decision thresholds when interpreting brief remote speech samples and during in-person evaluations. Similarly, confidence ratings differed substantially between raters and did not consistently protect against misclassification, indicating that clinician confidence should not be interpreted as a uniform proxy for diagnostic accuracy.

Finally, the composition of the sample also warrants consideration. Although the heterogeneous clinical cohort allowed MST to be evaluated across a range of neurologic and abnormal speech presentations, some diagnostic categories had relatively few participants. In particular, the dysphonia-only group was small, limiting conclusions about the ability of MST to distinguish isolated voice abnormalities from speech disorders. Studies with greater representation across neurologic diagnoses, motor speech disorder subtypes, and severity levels will be important for determining whether performance differs systematically across clinical populations. Future work should evaluate MST prospectively in independent clinical samples and determine whether standardized training or decision-support tools, such as explicit classification criteria and standardized scoring guidelines can reduce rater-related variability. The incorporation of automated speech analysis and artificial intelligence may also provide an opportunity to supplement clinician judgment by identifying speech features that are difficult to reliably detect from brief samples and, ultimately, improve the accuracy and consistency of remote motor speech screening.

## Conclusion

The present findings support the clinical utility of MST as a brief, remote evaluation approach to motor speech screening and triage. In approximately five minutes of recorded speech, MST demonstrated meaningful ability to distinguish normal from abnormal speech and to identify individuals with speech disorders, although performance and error patterns varied across raters. These findings suggest that the value of MST lies not in replacing the diagnostic depth of a comprehensive motor speech evaluation, but in helping determine who requires further evaluation and prioritizing those patients for a more comprehensive assessment by a speech-language pathologist.This distinction is particularly important in clinical environments where specialized speech-language pathology services are limited, and comprehensive evaluations may require substantial clinician time. A brief assessment that can be obtained remotely may provide an efficient first step for extending the reach of specialized motor speech expertise, identifying patients who warrant more comprehensive evaluation, and directing limited clinical resources toward those most in need. MST has the potential to support a scalable, tiered model of motor speech assessment in which remote screening facilitates timely access to comprehensive evaluation while preserving specialist resources for the diagnostic and treatment decisions that require them.

## Data Availability

All data produced in the present study are available upon reasonable request to the authors

